# Amphibian Collapses Exacerbated Malaria Outbreaks in Central America

**DOI:** 10.1101/2020.12.07.20245613

**Authors:** Michael R. Springborn, Joakim A. Weill, Karen R. Lips, Roberto Ibáñez, Aniruddha Ghosh

## Abstract

Ecosystems play an important role in supporting human welfare, including regulating the transmission of infectious diseases. Many of these services are not fully-appreciated due to complex environmental dynamics and lack of baseline data. Multicontinental amphibian decline due to the fungal pathogen *Batrachochytrium dendrobatidis* (Bd) provides a stark example. Even though amphibians are known to affect natural food webs—including mosquitoes that transmit human diseases—the human health impacts connected to their massive decline have received little attention. Here we show a causal link between a wave of Bd-driven collapse of amphibians in Central America and increased human malaria incidence. At the canton-level in Costa Rica and district-level in Panama, expected malaria incidence increased for eight years subsequent to amphibian losses, peaking at an additional 1.0 cases per 1,000 population (CPK). The increase is substantial in comparison to annual incidence levels from outbreaks in these countries, which peaked at 1.1-1.5 CPK during our period of study from 1976-2016. This pattern holds across multiple alternative approaches to the estimation model. This previously unidentified impact of biodiversity loss illustrates the often hidden human welfare costs of conservation failures. These findings also show the importance of mitigating international trade-driven spread of similar emergent pathogens like *Batrachochytrium salamandrivorans*.

**Significance Statement:** Despite substantial multicontinental collapses in amphibian populations from spread of the fungal pathogen *Batrachochytrium dendrobatidis* (Bd), the implications for humans have not been systematically studied. Amphibians are known to affect food webs, including mosquitoes that serve as a vector for the spread of disease. However, little is known about how their loss erodes ecosystem services, including the regulation of the transmission of infectious diseases. Using Central America as a case study, this study shows that Bd-driven amphibian loss led to a substantial increase in malaria incidence. The results highlight the often underappreciated social costs of biodiversity loss, including the potential stakes of ecosystem disruption from failing to stop spread of future novel pathogens.

---

Despite recent catastrophic, disease-driven loss of amphibians at a global scale, no broad implications for human welfare have been empirically demonstrated. Amphibians are not unique in this regard. While broad biodiversity loss impedes ecosystem functioning and the social benefits that follow, specific consequences of such change often go unnoticed (1). For amphibians, the spread of *Batrachochytrium dendrobatidis* (Bd)—an extremely virulent fungal pathogen responsible for massive worldwide die-offs (2)—has arguably caused “the greatest recorded loss of biodiversity attributable to a disease” (3). Empirical reckoning with implications for human welfare is essential for informed mitigation of ongoing impacts and—perhaps more importantly—sufficiently motivating investment to avoid repeating such disasters. For example, newer related pathogens like *Batrachochytrium salamandrivorans* similarly threaten to invade through the global movement of goods and people, repeating the cycle (4).

We take advantage of a natural experiment to provide what is to our knowledge the first causal evidence of a negative human health impact of widespread amphibian loss, namely through increased malaria incidence. The emergent One Health approach emphasizes ties between people, animals, plants, and environment, for example human-mammal/bird connections in outbreaks of novel influenza and coronaviruses (5). Less attention has been paid to connections between amphibians and human health. Over the past few decades in Central America biologists have tracked an invasion wave of Bd, which causes chytridiomycosis and has decimated amphibians. Loss of these species is known to affect ecosystem functions and natural food webs, with the potential to increase insect abundance, including mosquitoes capable of transmitting human diseases (6–10). This Bd wave travelled from west to east across Costa Rica from the early 1980s to the mid-1990s and then continued across Panama through the 2000s (11). Following this rolling collapse of amphibian populations, both countries experienced large increases in malaria cases. Fig. 1 shows annual total malaria cases in Costa Rica and Panama for the time span of our analysis. While the ordering and timing of peaks in the two countries are consistent with a lagged impact of amphibian decline, this correlation does not establish causality on its own.

**Fig. 1.**
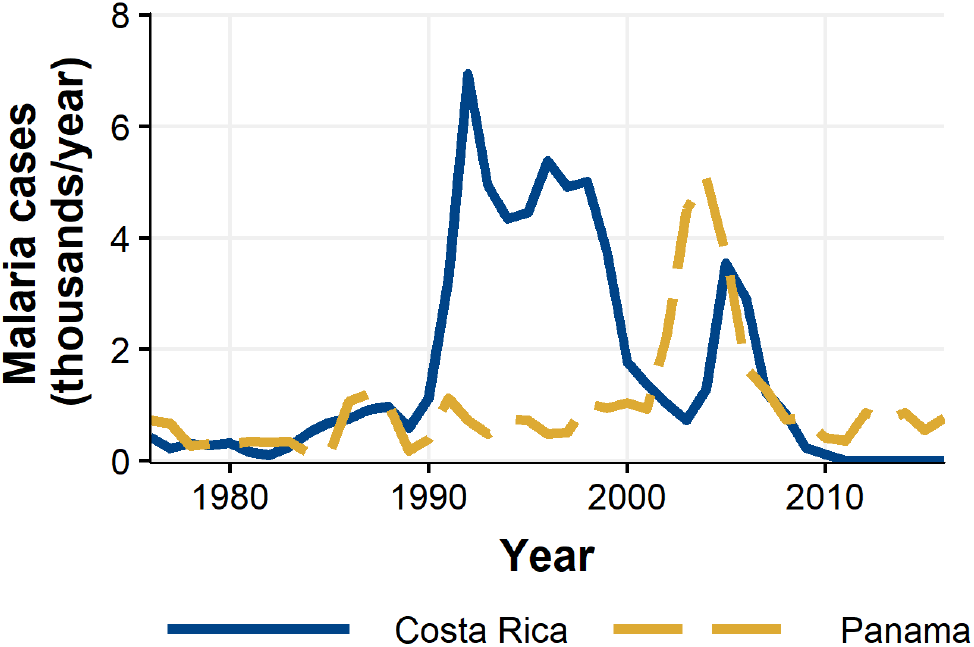
Annual total malaria cases from 1976-2016 for Costa Rica and Panama.

The global burden of malaria in 2018 includes an estimated 228 million cases and 405,000 deaths, largely in sub-Saharan Africa and India (12). Multiple overlapping social and environmental drivers have been proposed to explain instances of elevated malaria incidence. These include weather patterns, deforestation, human migration, and anti-malaria program problems (13, 14). Deforestation in particular has received increased attention in recent years and is hypothesized to operate through changes to the physical environment, malarial mosquito biology, and human exposure (15). While most have found that deforestation is associated with increased malaria incidence (16), this result does not hold across all regions and study designs (17). However, linkages between malarial dynamics and ecosystem disruption by invasive species has not been previously studied, aside from well-known linkages to invasive mosquito vectors.

## Methods

We use a multiple regression model to estimate the causal impact of Bd-driven amphibian decline on malaria incidence at the canton level in Costa Rica and distrito level in Panama, hereafter referred to as “county level”. We exploit variation in outcomes for units (counties) that experienced “treatment” (amphibian decline) at different times, which is a difference-in-difference, event-study design (18). This approach takes advantage of the staggered treatment of counties, leveraging differences in malaria outcomes over time between administrative units that have and have not been treated with Bd.

To estimate the linear regression model, we constructed a panel dataset spanning 41 years (1976-2016) at the county level in Costa Rica and Panama, as detailed in the SI Appendix. The outcome variable is malaria incidence (number of cases per 1,000 population). The central driver of interest is the Bd-driven date of decline (DoD) of amphibians in each county (when the edge of the county is first reached). We used field observations of the DoD at several sites across the two countries to estimate the DoD for each county using a spatial spread model on a grid overlaying the region (see SI Appendix). Fig. 2 shows the estimated DoD for each county included in our preferred specification. We excluded a small fraction of counties (indicated in Fig. 2) where precise DoD values were not available (see SI Appendix). The pattern shows a west-to-east wave spreading from the northwestern border of Costa Rica around 1980 to the Panama Canal region by 2010.

**Fig. 2.**
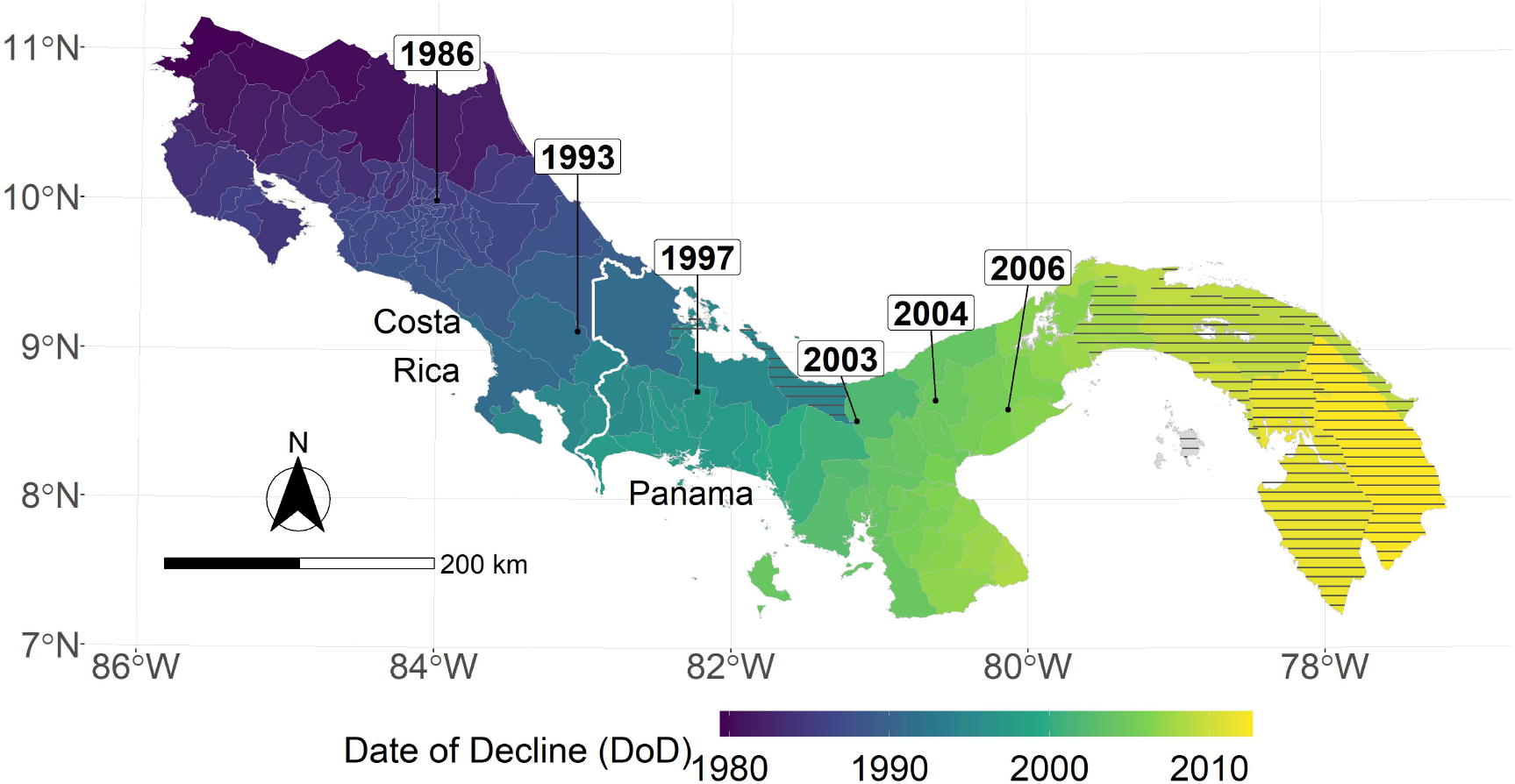
Date of Bd-driven amphibian decline (DoD) in Costa Rica and Panama. Observed DoD points are directly labeled with years. Color shading indicates county-level earliest DoD, estimated using a spatial spread process model. Hashing indicates counties excluded in the preferred specification.

For the regression model we use an event-study framework to characterize the impact of Bd-driven amphibian decline on per capita incidence of malaria, while controlling for other potential drivers. This approach is standard in the econometric literature in cases where multiple units, such as states or counties, receive the same “treatment” at different times (19–21). We specified the model as

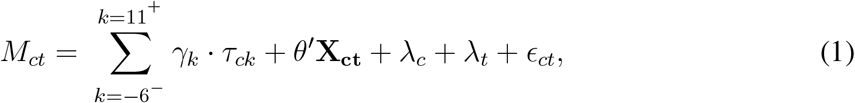

where *M*_*ct*_ is malaria cases per thousand inhabitants in county *c* and year *t*. The Bd-driven date of decline for county *c* is given by *DoD*_*c*_, where the unit is year. Also, at time *t* the number of years *relative* to this event is given by *K*_*ct*_ = *t* − *DoD*_*c*_. For example, if county *c* is “treated” by the arrival of Bd-driven amphibian decline at the start of 1990, then at *t* = 1992, the year relative to the event is *K*_*c*,1992_ = 1992 − 1990 = 2; county *c* has completed two years of treatment and is entering its third. Our main regressor of interest is *τ*_*ck*_, which is a dummy variable equal to one if county *c* is *k* years away from the initial treatment event: *τ*_*ck*_ = 𝟙{*K*_*ct*_ = *k*}. We focus on the five relative years before the event and 10 years following: *k* ∈ {−5, −4, …9, 10}. We also included a single dummy for all relative years before this, denoted by *k* = −6^−^ and another for all relative years after, *k* = 11^+^. Allowing the coefficient *γ*_*k*_ to vary for each relative year (*K*_*ct*_) in this way facilitates flexible and dynamic treatment effects. Because we imposed *γ*_−1_ = 0 to serve as the baseline, the remaining coefficients *γ*_*k*_ are interpreted as effects relative to the year *k* = −1, the year just before the DoD.

A vector of time-varying, county-level control variables is given by **X**_**ct**_. These include three annual weather measures—total precipitation and temperature extremes (minimum and maximum)—following previous analysis of malaria dynamics in Panama (13). We also consider annual land cover variables (share of tree cover, non-tree cover and bare ground) to account for the role of deforestation.

The regression model in Equation 1 also includes county fixed-effects (individual county dummies *λ*_*c*_) to control for differences between spatial units that are constant over time (e.g. elevation) as well as year fixed effects (individual year dummies *λ*_*t*_) to control for any shocks that affect malaria prevalence in all counties in a given year. Rounding out the model, *ϵ*_*ct*_ is a county-level error term.

The regression coefficients of interest, 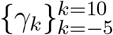, measure the relationship between the number of malaria cases per thousand inhabitants and the timing of the decline of amphibian populations in each county, conditional on the covariates. These estimates can be interpreted as causal as long as there are no omitted time-varying, county-level variables that both (1) impact malaria prevalence, and (2) are correlated with the wave of Bd-driven amphibian decline from west to east in our time frame. We argue that it is extremely unlikely that there exists such a variable that would satisfy both conditions, especially the second.^i^ This assumption would be violated for instance if each county received medical funding in a way that was systematically correlated with the decline of amphibian populations in these counties. However, such omitted systematic correlation is very unlikely.

## Results

### Effect of Amphibian Decline on Malaria

In Fig. 3 we plot the coefficients for the year relative to amphibian decline along with 90% confidence intervals for the preferred regression model. A crucial validity test of our event study framework is to confirm the absence of a pre-trend, i.e. a directional trend in the effect on malaria cases of the year relative to amphibian decline before treatment at year 0. Simply put, we would not expect to see systematic movement in malaria incidence in years before amphibian decline begins. We confirmed lack of such a pre-trend: none of the coefficients for *k* < 0 are significantly different from zero.

**Fig. 3.**
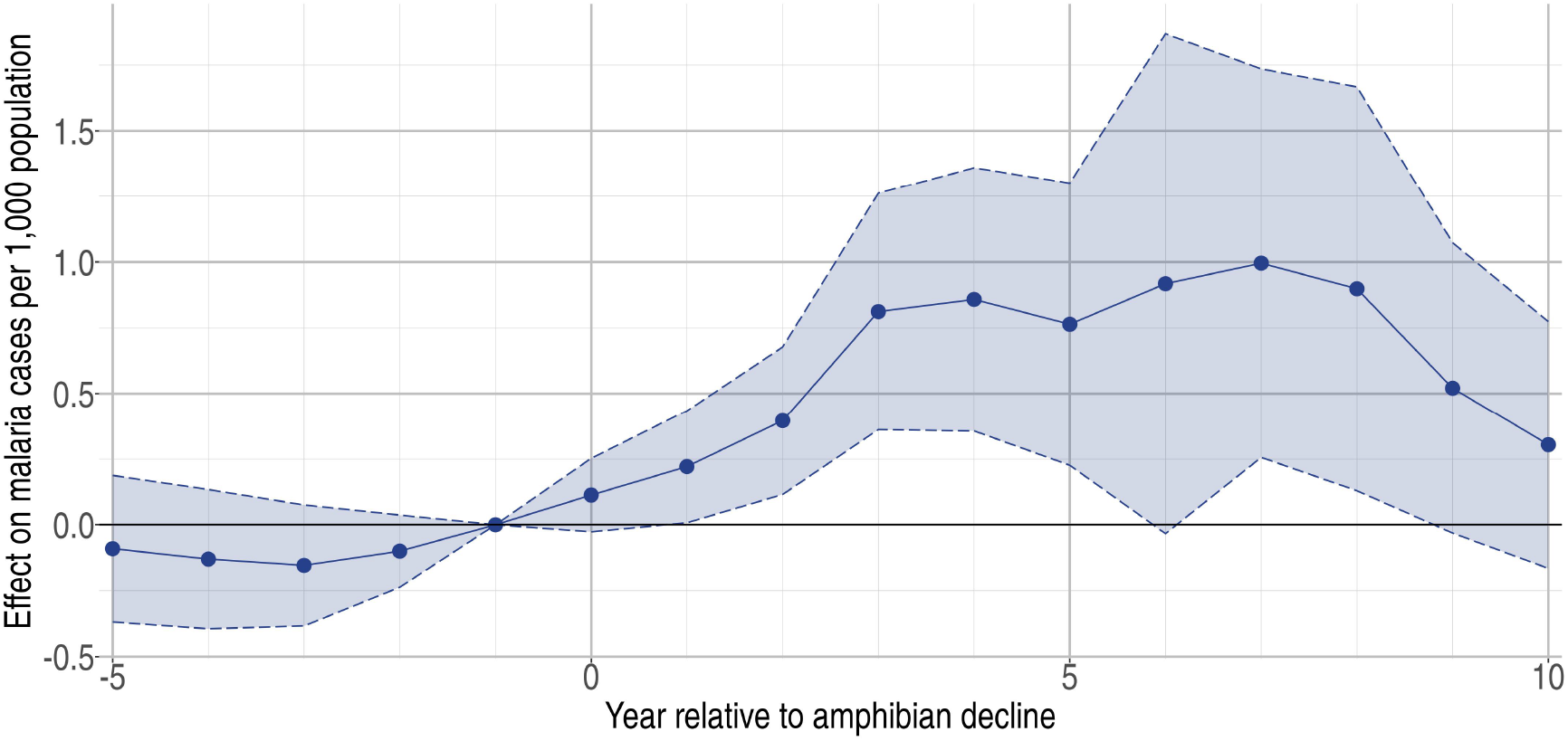
Estimated effect on malaria cases per 1,000 population (vertical axis) of year *k* (horizontal axis) relative to Bd-driven date of amphibian decline (DoD). Shading represents 90% confidence intervals.

Overall, we estimated a significant increase in malaria cases due to the onset of amphibian decline, an effect that starts gradually, plateaus after 3 years, and starts to attenuate after 8 years. The first year of amphibian decline (*k* = 0) reflects partial treatment for most counties since Bd-saturation of a county takes a median of 1.1 years and spread may arrive anytime in a calendar year. Starting in year *k* = 1, amphibian decline is associated with a statistically significant increase in malaria cases. We estimate that this average effect reaches a relative plateau by year *k* = 3 and stays relatively constant for six years. For one year in this range (*k* = 6) the effect is not significantly different from zero. This is not due to a decline in the coefficient but rather to an increase in the standard error due to an increase in residuals, i.e., additional noise. Starting in year *k* = 9 the average effect begins to attenuate and is no longer significantly different from zero.

For perspective on the relative magnitude of this Bd-driven effect, peak cases per 1,000 population reached approximately 1.1 for Panama (2002-2007) and 1.5 for Costa Rica (1991-2001). For the six years our estimated effect of amphibian decline is at its highest, the annual expected increase in malaria ranges from 0.76-1.0 additional cases per 1,000 population. This represents a substantial share of cases overall.

### Robustness Checks

In Table 1 we present the full set of regression estimates for the preferred specification (discussed above) in column 1, alongside estimates for three alternative specifications (columns 2-4) to check robustness (discussed further below). In the table, regression coefficients are presented with standard errors in parentheses, which are clustered at the county level. We clustered due to the sampling design (we are inferring something about the larger population based on data sampled at the county level) and our quasi-experimental design (“treatment” occurs at the county level). Overall, we found that our key qualitative results discussed above hold across an array of robustness checks.

**Table 1.** Estimates for the regression model specified in Equation for the preferred specification (column 1) and alternatives for robustness checks. Standard errors (clustered at the county level) are presented in parentheses.

|  | *Dependent variable: malaria cases per 1,000 population* |  |  |  |
| --- | --- | --- | --- | --- |
|  | Preferred specification | All observations | Average date of decline | Weighted regression |
|  | (1) | (2) | (3) | (4) |
| Tree cover | −2.556** (1.171) | −2.891** (1.174) | −2.551** (1.166) | −2.005** (0.869) |
| Bare ground | −1.823 (1.379) | −2.330* (1.394) | −1.737 (1.312) | −1.222* (0.706) |
| Precipitation | 0.002 (0.002) | 0.002 (0.002) | 0.003 (0.002) | 0.001 (0.001) |
| Min. Temp. | 0.295 (0.186) | 0.071 (0.248) | 0.318* (0.190) | 0.315 (0.212) |
| Max. Temp. | 0.222 (0.181) | 0.363 (0.235) | 0.240 (0.187) | 0.057 (0.111) |
| $\hat{\gamma}_{-6-}$ | −0.448*** (0.159) | −0.431** (0.176) | −0.546*** (0.196) | −0.323*** (0.106) |
| $\hat{\gamma}_{-5}$ | −0.091 (0.169) | −0.043 (0.202) | −0.190 (0.185) | −0.095 (0.104) |
| $\hat{\gamma}_{-4}$ | −0.130 (0.160) | −0.150 (0.179) | −0.218 (0.152) | −0.076 (0.095) |
| $\hat{\gamma}_{-3}$ | −0.154 (0.139) | −0.286* (0.168) | −0.214 (0.135) | −0.107 (0.086) |
| $\hat{\gamma}_{-2}$ | −0.100 (0.083) | −0.172 (0.119) | −0.199** (0.086) | −0.036 (0.062) |
| $\hat{\gamma}_0$ | 0.113 (0.085) | 0.109 (0.083) | 0.209** (0.097) | 0.053 (0.049) |
| $\hat{\gamma}_1$ | 0.221* (0.130) | 0.141 (0.140) | 0.444** (0.213) | 0.109 (0.074) |
| $\hat{\gamma}_2$ | 0.396** (0.171) | 0.277 (0.173) | 0.716*** (0.272) | 0.340** (0.134) |
| $\hat{\gamma}_3$ | 0.812*** (0.274) | 0.750*** (0.264) | 0.708** (0.289) | 0.762** (0.318) |
| $\hat{\gamma}_4$ | 0.858*** (0.305) | 0.866*** (0.293) | 0.652* (0.359) | 0.842** (0.383) |
| $\hat{\gamma}_5$ | 0.764** (0.327) | 0.870*** (0.320) | 1.023 (0.682) | 0.663** (0.267) |
| $\hat{\gamma}_6$ | 0.917 (0.578) | 0.937* (0.563) | 0.880* (0.510) | 0.635 (0.386) |
| $\hat{\gamma}_7$ | 0.995** (0.450) | 1.086** (0.447) | 0.553 (0.425) | 0.832** (0.361) |
| $\hat{\gamma}_8$ | 0.898* (0.467) | 1.052** (0.465) | 0.257 (0.324) | 0.664** (0.331) |
| $\hat{\gamma}_9$ | 0.521 (0.336) | 0.962** (0.447) | 0.158 (0.316) | 0.413* (0.245) |
| $\hat{\gamma}_{10}$ | 0.304 (0.285) | 0.649* (0.349) | 0.087 (0.299) | 0.253 (0.224) |
| $\hat{\gamma}_{11+}$ | 0.645 (0.401) | 0.676* (0.398) | 0.455 (0.418) | 0.532* (0.300) |
| Year FE | Yes | Yes | Yes | Yes |
| County FE | Yes | Yes | Yes | Yes |
| Observations | 5,576 | 5,904 | 5,576 | 5,576 |
| Adjusted R<sup>2</sup> | 0.301 | 0.311 | 0.302 | 0.310 |

Note:
\*p<0.1; \*\*p<0.05; \*\*\*p<0.01

In the table, our independent variables (rows) start with two ground cover measures and three weather measures. Next are the key coefficients of interest, 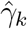, representing the estimated effect of relative years before a county’s DoD (*k* < 0) and after (*k* ≥ 0). These coefficients for our preferred specification for *k* = −5, −4, …, 10 are plotted in Fig. 2 and discussed above. We excluded *k* = −1 so that the rest of these coefficients are interpreted as effects relative to this year just before a county’s DoD. For our first alternative specification in column (2) we augmented the data set with regions of Panama excluded in our preferred specification due to data limitations as described in the SI Appendix.^ii^ In column (3) we considered an alternative rule for converting the raster of DoD levels at the pixel-level to the county-level: instead of the using the minimum date reflecting initial arrival to the county border, we considered the average DoD for the county. In column (4) we conducted weighted least squares regression where the weights were given by county-level population. This heightened emphasis on observations from high-population counties is motivated by the conjecture that malaria incidence measures from such counties are less subject to measurement error given the larger number of “samples” available.

For our key coefficients of interest, under our preferred specification none of the pre-DoD coefficients(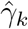 for *k* ∈ ⟦−5, −2⟧) are significantly different from zero, i.e. we fail to find a significant pre-trend in malaria in the five years leading up to the DoD. In event study frameworks like the one used here, such a lack of a pre-trend is one critical check for model validity. In subsequent relative years after Bd-driven amphibian decline, the coefficients are positive, significantly so for 1 ≤ *k* ≤ 8, with the exception of one year (*k* = 6). Lack of significance in this year appears to stem from an uptick in noise—the coefficient is within the range of surrounding years while the standard error is elevated.

We found that the same general pattern holds across all specifications: we fail to find a pre-trend before the DoD and then find a block of significant positive effects on malaria subsequent to the DoD. This pattern shifts earlier in relative years under specification (3) as we would expect. In this specification *k* = 0 does not indicate the beginning of treatment but rather a time when approximately half of the county has already been treated. In this case the omitted year *k* = −1 (for which the effect is assumed to be zero) includes partially treated counties, skewing the baseline and leading to a likely spurious significant negative coefficient for 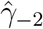. This justifies focus on our preferred specification in which all treatment begins at *k* = 0, aiding consistent interpretation of the coefficients.

In results presented in the SI Appendix, we also examined the sensitivity of estimates to an alternative method used for estimating the gridded DoD values, specifically a thin-plate spline (TPS) method for direct interpolation of the DoD data over space. Although this method ignores the implications for spread of Central America’s irregular coastline, the results are also broadly consistent with our preferred specification.

### Additional Drivers of Malaria

We also found that decreasing tree cover is associated with a statistically significant increase in malaria cases under all specifications (see first row of Table 1) in keeping with the majority of findings of previous studies. A one standard deviation decrease in tree cover (0.05) is associated with an increase of 0.13 in the number of cases of malaria per 1,000 inhabitants. This is about one-eighth the magnitude of the estimated amphibian decline impact at its peak. Bare ground also has a negative effect on cases, though this effect is not consistently significant. Non-tree vegetation, the third land cover type, was excluded from the regression because all three types sum to one for each county; the excluded type is perfectly multicollinear with the sum of the two included measures.

For weather variables, increasing precipitation and higher temperatures were associated with additional cases, though typically not significantly so (except for minimum temperature in one specification (3)).

## Discussion

Overall we provide novel causal evidence that pathogen-driven amphibian decline can play a significant role in increasing incidence of vector-borne disease. Our results also contribute to a nascent but growing literature identifying indirect and previously unknown impacts of invasive species on human health (22–24). If scientists and decision makers fail to reckon with the ramifications of such past events, they also risk failing to fully motivate protection against new calamities, like international spread of an emergent and closely related pathogen *Batrachochytrium salamandrivorans* through incompletely regulated live species trade (4).

While results were robust to several alternative specifications, we were unable to examine whether outcomes for malaria held for other diseases. Showing that our results held for other vector-borne diseases (e.g. dengue and leishmaniasis) would have provided additional support for the mechanism we propose. Showing that our results failed to hold for non-vector-borne illnesses like influenza would have provided additional support for the argument that the effect we identify is specific to vector-borne diseases and not a general disease effect. We attempted to obtain these disease data sets from the national ministries of health in both countries but they were not available for our period of study at the county-level needed.

From the data we were able to obtain from the Panamanian Ministry of Health, we found that the national-level time series for both dengue and leishmaniasis (vector-borne diseases) were consistent with the spike observed in malaria cases for 2002-2007 shown in Fig. 1. Relative to a baseline from the preceding ten years, average annual leishmaniasis cases were 22% higher for 2002-2007. For dengue the increase relative to the previous eight years (all available) was 36%. When we extended the window of potential impact to 2002-2011, average annual cases increased relative to baselines by 23% and 61%, respectively.

A puzzle for future study is why the estimated effect (of Bd-driven amphibian loss) attenuates, here after approximately 8 years. One plausible explanation is an increased malaria prevention program response to an observed uptick in malaria cases, e.g. increased investment in control measures like insecticide application. In the SI Appendix we discuss indicators of national malaria prevention actions (total funding and number of houses sprayed for mosquitoes) for our period of study for both countries (25). Total funding dynamics in both countries show increased (though sometimes uneven) investment in malaria prevention following national outbreaks, which would plausibly serve to suppress cases over time. While the evidence is suggestive we were not able to include these time series in our regression model since they are not available at the county level and such investment is endogenous with malaria cases (the outcome variable of interest).

## Data Availability

Land cover data were published by Song et al. (2018), Weather data were provided by Hijmans (2019), Amphibian collapse data were provided by Karen Lips, Malaria records were obtained from the Ministries of Health in Costa Rica and Panama. All data will be made available on a Github repository.

## Footnotes

^i^Formally, this identifying assumption is 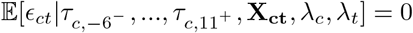.

^ii^Excluded regions included eastern Panama and the re-aggregated district “Bocas del Toro”.

## Acknowledgments

We thank L. Fonseca for research assistance, R. Hijmans for spatial modeling advice, and the following individuals for assistance obtaining malaria data: M. Piaggio (Environment for Development); T. and R. Castro (Costa Rica Ministry of Health); and D. Batista, E. Benavides, M. Lasso and L. García (Panama Ministry of Health). We also thank D. Rodríguez and L. Moreno (Panama Ministry of Health) for providing the leishmaniasis and dengue data.

## Contributions

Conceptualization (K.R.L., M.R.S. and J.A.W.), Data curation (all), Methodology and Formal Analysis (M.R.S. and J.A.W.), Funding acquisition (M.R.S.), Investigation (all), Project administration (M.R.S.), Visualization (A.G., M.R.S and J.A.W.), Writing original draft (M.R.S. and J.A.W.), Writing review and editing (all).

## Supplementary Information

### Data Overview

Below we describe data sources and methods for constructing our panel data set spanning each year from 1976-2016 at the canton-level in Costa Rica and distrito-level in Panama, hereafter referred to as “county-level”. This core data set includes variables presented in Table S1 as well as the Bd-driven date of amphibian decline (DoD) over space as presented in Fig. 2. Because Panama experienced multiple distrito boundary changes during our study period, we aggregated certain distritos for spatial consistency. We omitted a small set of distritos and regions that present aggregation problems or are islands. These steps are described in detail further below. After these adjustments, our main data set includes 136 counties (81 in Costa Rica and 55 in Panama).

**Table S1.**
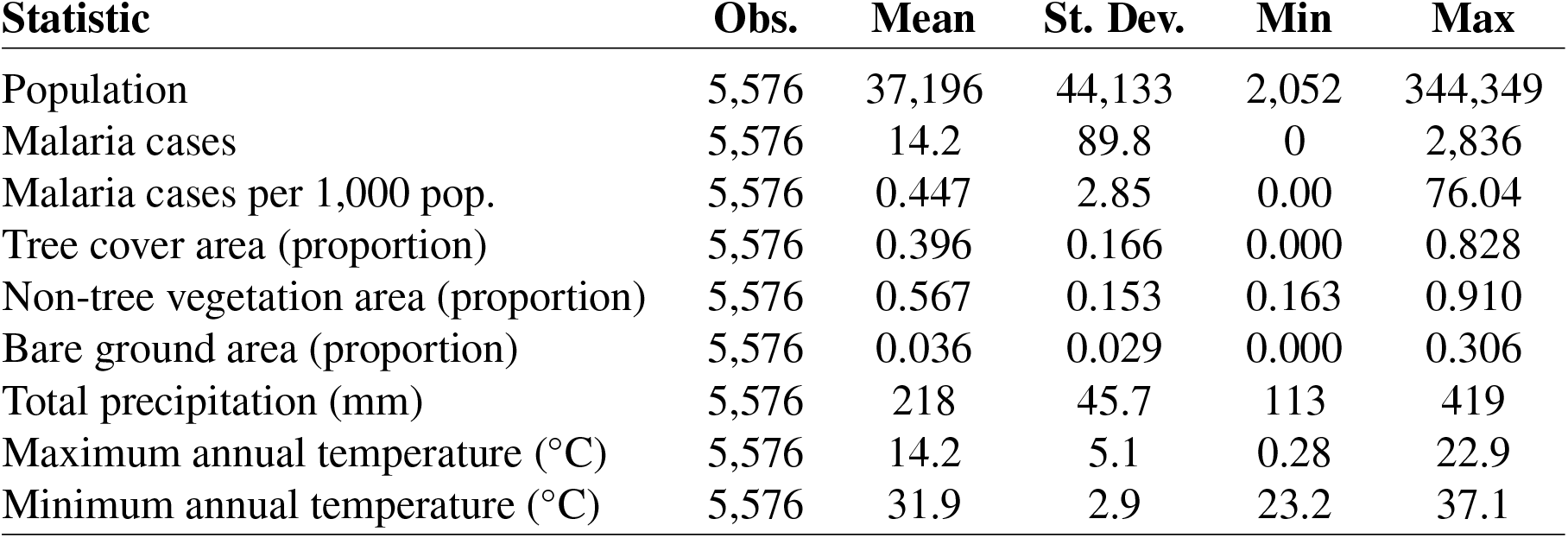
Summary statistics for annual (1976-2016), county-level variables used in the baseline regression model.

### Malaria Cases and Population

Annual malaria cases were digitized from administrative records obtained from the Ministries of Health in Costa Rica and Panama. While data do not distinguish between *Plasmodium vivax* and *Plasmodium falciparum* the vast majority of cases in this region and period are due to the former.^i^ We compiled county-level population data from administrative records provided by the National Institute of Statistics and Census in Costa Rica and in Panama.^ii^ For Panama, distrito-level population data was available at decadal intervals from 1970-2010. We used cubic hermite splines to interpolate annual measures. For Costa Rica, canton-level population was available for all years from 1976-2016, except 1978-1981. Given the small, four-year span to fill in, we used linear interpolation between available data in 1977 and 1982.

### Land Cover

For measures of annual land cover for 1982-2016 we use used a dataset with global coverage at a spatial resolution of 0.05° × 0.05° produced by Song et al. (1). The measures include the share covered by tree canopy (taller than 5 meters), short vegetation (shorter than 5 meters), and bare ground. These dataset were derived from the Advanced Very High Resolution Radiometer (AVHRR) version 4 Long Term Data Record and represent the annual land cover status at peak growing season for each pixel (1). From this annual raster data set, we aggregated the measures to the county level.

Satellite data from the first six years in our study period (1976-1981) is much more limited over time, over space, and in sensor. Thus, we did not use remote sensing products for these years. Instead we obtained national measures of the annual change in tree canopy (described below) and assumed that any reported rate of change at the national level is the same across counties within each country. Because such downscaling can generate time series that are much smoother than years actually observed, we added noise to the downscaled values. To do so, we computed for each county the standard deviation of tree cover between 1981 and 2001. We then generated random draws from centered normal distributions with the previously estimated standard deviations, and we added these draws to the interpolated values of tree cover between 1976 and 1981. We also assumed that for the remaining land use that is not tree canopy, the relative split between short vegetation and bare ground is the same in each county as in 1982.

National measures of the annual change in tree canopy cover from 1976-1981 were established as follows. For Costa Rica, we used the national annual forest cover percentage for 1963-1994 as estimated by the Food and Agriculture Organization (FAO) of the United Nations (2). From 1976-1981, FAO forest cover estimates in Costa Rica drop by a constant rate of 1.49 percentage points per year. FAO estimates for this period are not available for Panama. We used estimates from the academic literature reporting that, from 1976-1980, Panama lost an average of 31,000 hectares annually (3), which is equivalent to 0.42 percentage points per year. We assumed this rate also holds for one additional year (through 1981).

To provide a sense of the magnitude of the impact of deforestation on malaria incidence, we first identified a plausible change in the tree cover variable. We followed the approach of (4) and residualized the tree cover variable with respect to both county and year fixed effects. The residuals have a standard deviation of 0.05 during our entire panel. The standard deviation is very similar (0.04) if we instead restrict the panel to the period of malaria outbreaks in each country. We then obtained the impact of tree cover on malaria by multiplying this plausible shift (0.05) by the regression coefficient, as reported in the main text.

### Weather Variables

Estimates of three monthly weather variables—total precipitation and average daily minimum and maximum temperature—at a high level of spatial resolution (0.04°, approximately 4.4 km) for 1976-2016 were obtained from Hijmans (5).^iii^ From this monthly, gridded data set we constructed annual statistics for each county. For precipitation, we computed the average (over grid cells) of the monthly county-level total, which we then aggregated to the yearly total. For minimum (maximum) temperature, we obtained the monthly minimum (maximum) level across grid cells in each county and then selected the minimum (maximum) level over all months in each year.

### Declines in Amphibian Populations

The central explanatory variable of interest in our regression model is the Bd-driven date of decline of amphibian populations (DoD) in each county. To estimate these dates, we first collected all available observed DoD values reported in the ecological literature (6–10). Each of these observations specifies the date at which Bd-driven decline first occurred at given set of geographic coordinates. These observations—directly labeled with years in Fig. 2—indicate a consistent wave of Bd spread eastward from western Costa Rica through Panama.

We leveraged these spatiotemporal DoD data points to estimate dates for each county. To do so, we estimated a pixel-to-pixel Bd spread model that minimizes the sum of squared residuals (SSR) between the observed and fitted DoD values. The parameters selected to optimize the fit are the rates of spread at each of the DoD data points. For any given set of these rates, we extrapolated rates of spread across all pixels using ordinary Kriging. We then used these pixel-specific spread rates to simulate Bd spread from western Costa Rica through Panama. While a pixel-to-pixel spread model is computationally costly to repeatedly evaluate (e.g. for alternative parameter vectors in an estimation routine), it facilitates more realistic spread along irregular coastlines and the nonlinear isthmus of Costa Rica and Panama. This modeled spread results in a fitted DoD for every pixel, including those for which we have observed DoDs to calculate the residuals for minimization.

Because evaluation of the residuals resulting from a given set of parameter estimates is not a direct function calculation, we used an iterative algorithm for optimization. We started with a guess for the parameter vector of spread rates, which is informed by the observed physical and temporal distances between data points.^iv^ Then, in each iteration of the algorithm, one of the parameters was randomly selected to be perturbed, both up and down, by a fixed adjustment factor (e.g. 5%). Then, from the three candidate models (unperturbed, selected parameter adjusted up, selected parameter adjusted down) the one with the lowest SSR is retained and used as the basis for the next iteration. When improvements in the SSR cease, the adjustment factor is halved (e.g. from 5% to 2.5%) to further refine the solution. We performed 200 iterations of the algorithm since after 175 iterations, no further improvements in SSR were typically achieved. To guard against this stochastic algorithm identifying a local and not a global optimum, the best-performing model is selected from the set resulting from running the algorithm 100 times. Overall, the final spread model fits the observed data well—the absolute residuals (difference between observed and fitted DoD values) have an average of 0.15 years and a maximum of 0.23 years.

The result of the final solution is a raster map of DoD values. We converted these gridded values to a value for each county by taking the earliest DoD within each county, presented as the shaded values in Fig. 2. We also considered the average DoD value in each county as an alternative specification in our regression results.

As another robustness check, we considered a wholly different method for estimating county DoD values: directly fitting a thin plate spline (TPS) function for the DoD continuously over space. A TPS is two-dimensional extension of a cubic spline. Formally, the TPS *f* (**x**; *ω*) is a function of the underlying variables **x**—in our case longitude and latitude coordinates—and a parameter vector *ω* selected to miminize a combination of the SSR and a penalty for increasing curvature, quantified by the integral of squared second order derivatives:

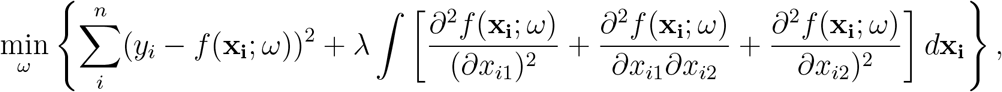

where *x*_*i*1_ and *x*_*i*2_ are the longitude and latitude coordinates at point *i*, and *y*_*i*_ is the observed DoD at that point. *λ* is a parameter that penalizes the curvature. If *λ* = 0, any “amount” of curvature is allowed and the spline will perfectly match the data. If *λ* → ∞, no curvature is allowed and the optimization problem boils down to ordinary least squares.

In practice, *λ* can be fixed at a given value or selected through a cross-validation method. We selected *λ* using generalized cross validation and then further assessed the robustness of our results to values of *λ* ranging from its minimum to a point where the resulting output no longer changed. The result of the interpolation provides a smooth surface of DoD values, as shown in Fig. S1, which are then converted to county-level values in the same way as for the spread model described above.

The TPS approach is appealing in its simplicity as a one-step fitting procedure that directly extrapolates observed DoD values over space. However, the pattern in Fig. S1 illustrates a key drawback of this method: it ignores realities of irregular landscape shapes, in essence allowing for spread unimpeded across ocean, equivalently to land.

**Fig. S1.**
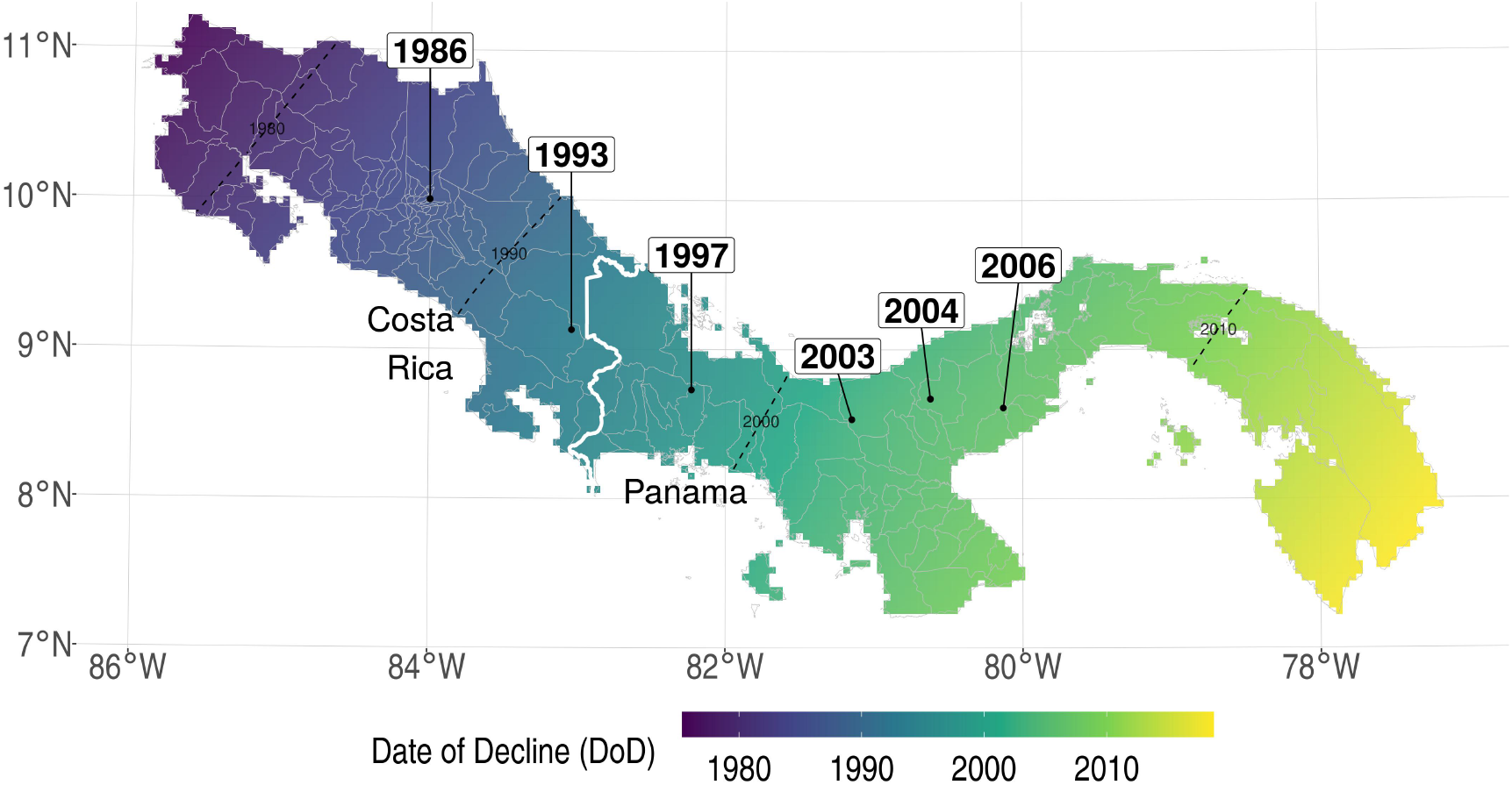
Bd-driven date of amphibian decline (DoD) in Costa Rica and Panama estimated with a thin plate spline with the curvature penalty selected using generalized cross validation (*λ* = 12.5). Observed DoD points are directly labeled with years. Color shading indicates estimated DoD values.

### Spatial Units

Our “county-level” spatial unit of analysis corresponds to cantons in Costa Rica and distritos in Panama. There have been several distrito boundary changes in Panama over the past few decades.^v^ To maintain consistent spatial units over time, we aggregated the data of distritos that experienced a split in the middle of our study period (1976-2016). This approach follows related previous analysis (11) and facilitates keeping the smallest spatial units for which we can unambiguously attribute population and malaria counts during the entire period.^vi^

We omitted from our preferred regression model estimation two distritos and one region that either present aggregation problems or are islands. We excluded island distritos (Balboa and Taboga) since we were concerned with land-based spread of Bd and were unable to rigorously predict if and when it arrived to offshore locations. On mainland Panama, we omit the relatively small Canal Zone due to aggregation problems. This used to be a U.S. special area which included a strip of land on both sides of the Panama Canal. This area fully returned to the Republic of Panama on December 31st, 1999, and was distributed among the Colón district, the Panamá district, the Chorrera district and the Arraiján district. We omit the Canal Zone since keeping it and ensuring consistent boundaries would require aggregation of four otherwise distinct districts.

Finally we omit two regions of Panama for which we are unable to rigorously predict the arrival of Bd. First, the re-aggregated district “Bocas del Toro” in northwestern Panama—folding in Bocas del Toro and Kusapín, which were split off in 1997—has a particularly problematic geography. It is extremely wide, spatially discontiguous and features an irregular coastline, such that the estimated delay between Bd-arrival at one tip and saturation to the other is a substantial outlier at 8.0 years. For counties in our preferred specification, this estimated time to saturation has a median of 1.1 years (standard deviation 1.3 years). Over 90% of counties are saturated in less than 3 years. Because saturation of the aggregated Bocas del Toro county takes more than 7 times the median duration, we exclude it from the preferred specification.

The second region we omit from our preferred specification encompasses eastern Panama (hashed region in Fig. 2) since it is relatively far away from the last DoD observation, and DoD predictions in this zone may be particularly imprecise.^vii^ In our discussion of robustness checks (further below), we found that key results are not sensitive to inclusion of Bocas del Toro and eastern Panama.

### DoD Model Robustness Check

As an additional robustness check, we examined the sensitivity of our results to the method used for estimating the DoD at the pixel level. For the results presented below we used the thinplate spline (TPS) method for directly estimating DoD from the data (as described above). We considered eight different implementations of TPS, which vary in the curvature penalty parameter *λ* (i.e. the inverse “flexibility” of the spline). We assessed a range from maximally flexible (*λ* = 0) through increasingly inflexible splines until results no longer vary. The resulting upper bound on this penalty parameter coincided with the level also selected using generalized cross validation (*λ* = 12.5). Across this set, there is very little variation in the key regression coefficient estimates, as shown in Fig. S2. The results are also broadly consistent with our preferred specification, with significant increases in malaria by year three after the DoD. One exception is that the lack of a pre-trend before the DoD (shown in the preferred specification) is not as clear here—some coefficients for relative years *k* < −2 are significantly different from zero. However, results from this TPS approach are not expected to be precise given that the interpolation method ignores realities of the irregular coastlines in Costa Rica and Panama (as discussed in the TPS description above) that are accounted for in our preferred approach.

**Fig. S2.**
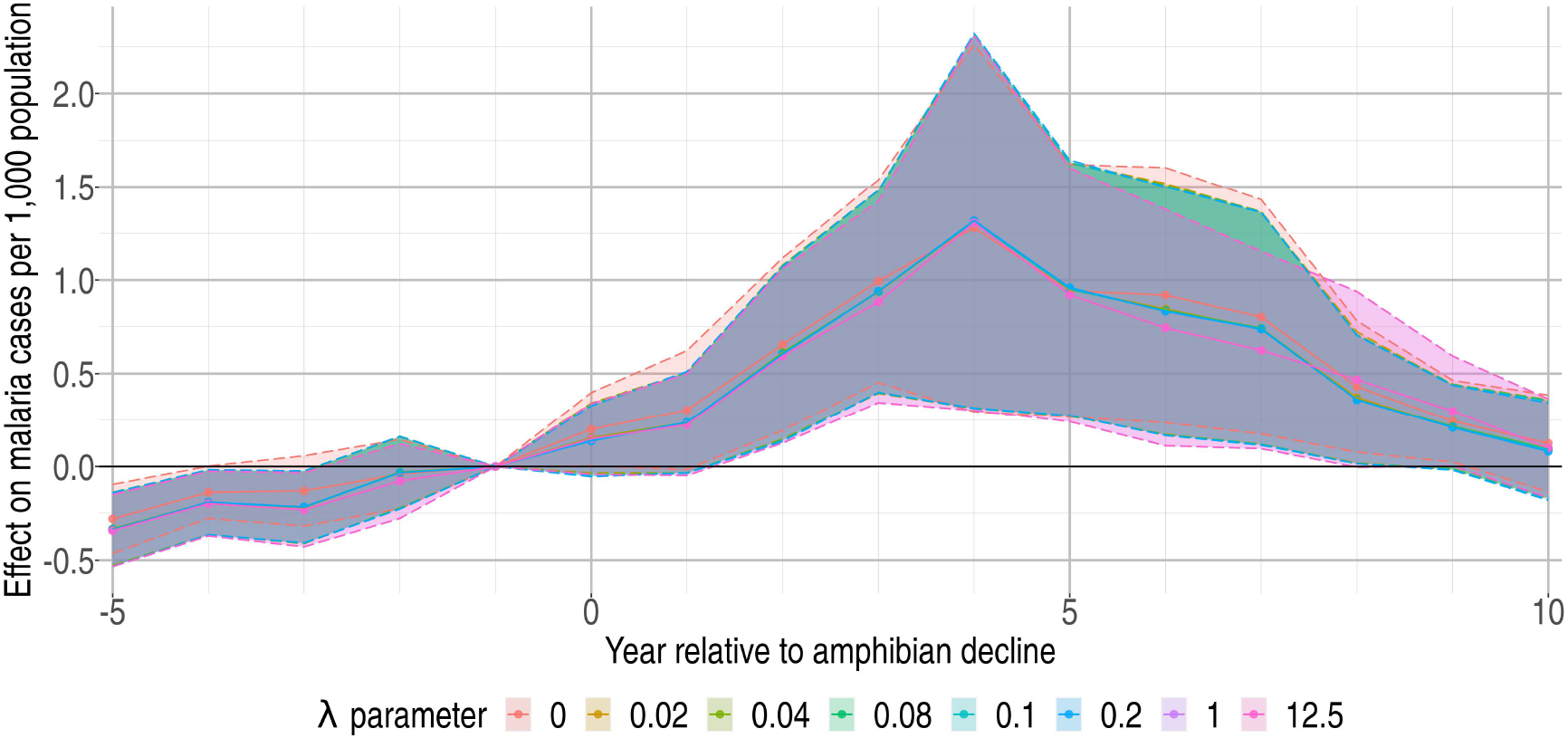
Estimated effect on malaria cases per 1,000 population (vertical axis) of the *k*-th year relative to the DoD or Bd-driven date of amphibian decline (horizontal axis) where DoD is estimated using a thinplate spline (TPS). Lines depict results under a range of TPS interpolations, which vary in the penalty parameter, *λ*. Shading represents 90% confidence intervals.

### National Malaria Prevention Actions

In main text Fig. 1 we show that during our period of study malaria cases spiked in Costa Rica 1991-2001 with a peak in 1992 and later spiked in Panama 2002-2007 with a peak in 2004. A second small spike occurred in Costa Rica in 2004-2007 with a peak in 2005. One possible factor in limiting these outbreaks is investment in malaria prevention programs. In Fig. S3 we show overall malaria prevention expenditures and number of households sprayed for mosquitoes in Costa Rica and Panama over the time range of our study from the Pan American Health Organization (PAHO).

In Costa Rica, relative to the 1980s, we see a substantial uptick in total funding (government and external) starting 1993 followed by massive increases in 1998 and again in 2008. For Costa Rica, these funding pulses are correlated first with the onset of the spike in cases and then eventual decline. While spraying houses for mosquitoes increases somewhat in the late 1990s, spraying ultimately decreases to fairly low levels. In Panama a similar dynamic is present though less consistent—a modest increase in malaria financing follows the pulse of cases in the early 2000s. However from 2008-2011 financing falls to the lowest levels since 1990 before accelerating to all time highs in 2012-2014. Spraying in particular is a plausible factor in reducing cases given a spike in spraying 2005-2010.

**Fig. S3.**
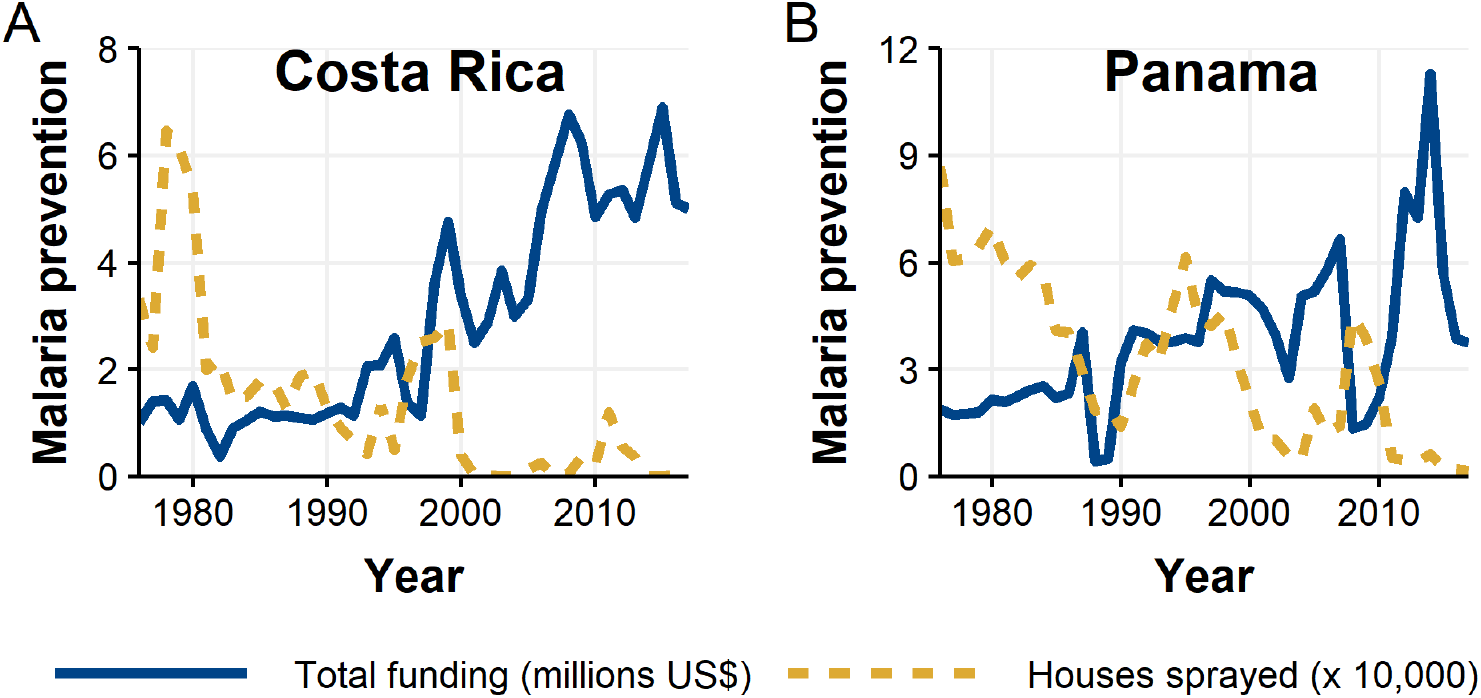
Annual total malaria prevention expenditures and houses sprayed for mosquitoes from 1976-2017 for Costa Rica (**A**) and Panama (**B**).

## Footnotes

i See Interactive Malaria Statistics by the Pan American Health Organization and World Health Organization, specifically ‘Cases by species type’, available at https://www.paho.org/data/index.php/en/mnu-topics/indicadores-malaria-en.html.

ii See records available at https://www.inec.cr/ and https://www.contraloria.gob.pa/INEC, respectively.

iii To construct these variables, Hijmans (5) obtained these monthly climate variables at a 0.5° spatial resolution from the Climate Research Unit data website (12, 13) and downscaled to the finer spatial resolution using the delta method (14).

iv An initial estimate of the rate of spread (meters/week) at each of the DoD data points is calculated as follows. At a given point *i*, we combined information on how fast spread reaches (A) *i* from the previous point *i* − 1 and (B) *i* + 1 from *i*. Considering points *i* and *i* − 1, let *t*_*i*_ represent the DoD time difference and let *d*_*i*_ represent the physical distance. The observed rate of spread from from *i* − 1 to *i*, or “approach rate” is given by *d*_*i*_*/t*_*i*_. We specified the rate of spread at point *i* as a weighted combination of the approach rate and departure rate, 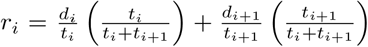. This form places greater weight on the approach rate if the spread process spends more time at this rate, relative to the departure rate (and vice versa). For the earliest and latest DoD observations, rate of spread is only available to one side of the point, which receives full weight (1). We used the rate of spread at the first point in Costa Rica to infer the date of arrival at the northwestern border of Costa Rica.

v Most notably, the Comarca Ngäbe Buglé was created in 1997 and led to the creation of eight new districts, carved out of previously existing districts. For instance, the district San Félix was split into San Félix and Mironó.

vi We aggregated distritos in the following way: Mironó was merged with San Félix, Nole Duima was merged with Remedios, Kusapín was merged with Bocas del Toro, Cémaco was merged with Pinogana, Sambú was merged with Chepigana, Müna is merged with Tolé, Besikó was merged with San Lorenzo, Kankintú was merged with Chiriquí Grande, Ñürüm and Cañazas were merged with Las Palmas, Mariato was merged with Montijo.

vii This includes the provinces of Panamá and Darién and the Comarca Guna Yala.

## References

[1] EF Zipkin, GV DiRenzo, JM Ray, S Rossman, KR Lips, Tropical snake diversity collapses after widespread amphibian loss. Science 367, 814–816 (2020).

[2] L Berger, et al., Chytridiomycosis causes amphibian mortality associated with population declines in the rain forests of Australia and Central America. Proceedings of the National Academy of Sciences of the United States of America 95, 9031–9036 (1998).

[3] BC Scheele, et al., Amphibian Fungal Panzootic Causes Catastrophic and Ongoing Loss of Biodiversity. Science 363, 1459–1463 (2019).

[4] TT Nguyen, T Van Nguyen T Ziegler, F Pasmans, A Martel, Trade in wild anurans vectors the urodelan pathogen batrachochytrium salamandrivorans into europe. Amphibia-Reptilia 38, 554–556 (2017).

[5] ME El Zowalaty, J. Jaärhult, From sars to covid-19: A previously unknown sars-cov-2 virus of pandemic potential infecting humans–call for a one health approach. One Health 9 (2020).

[6] G Bowatte, P Perera, G Senevirathne, S Meegaskumbura, M Meegaskumbura, Tadpoles as dengue mosquito (aedes aegypti) egg predators. Biological Control 67, 469–474 (2013).

[7] SE DuRant, WA Hopkins, Amphibian predation on larval mosquitoes. Canadian Journal of Zoology 86, 1159–1164 (2008).

[8] MJ Rubbo, JL Lanterman, RC Falco, TJ Daniels, The influence of amphibians on mosquitoes in seasonal pools: Can wetlands protection help to minimize disease risk? Wetlands 31, 799–804 (2011).

[9] G Bowatte, P Perera, G Senevirathne, S Meegaskumbura, M Meegaskumbura, Tadpoles as dengue mosquito (Aedes aegypti) egg predators. Biological Control 67, 469–474 (2013).

[10] DJ Hocking, KJB Abbitt, Amphibian Contributions to Ecosystem Services. Herpetological Conservation and Biology 9, 1–17 (2014).

[11] KR Lips, J Diffendorfer, JR Mendelson, MW Sears, Riding the wave: Reconciling the roles of disease and climate change in amphibian declines. PLoS Biology 6 (2008).

[12] World Health Organization, World Malaria Report 2019. (World Health Organization), (2019).

[13] LA Hurtado, L Caáceres, LF Chaves, JE Calzada, When climate change couples social neglect: malaria dynamics in panamaá. Emerging Microbes & Infections 3, 1–11 (2014).

[14] Grupo Teécnico Nacional de Enfermedades Vectoriales (Costa Rica), Plan de eliminacioón de la malaria en Costa Rica, 2015-2020 (https://www.binasss.sa.cr/planmalaria.pdf) (2015).

[15] JM Tucker Lima, A Vittor, S Rifai, D Valle, Does deforestation promote or inhibit malaria transmission in the Amazon? A systematic literature review and critical appraisal of cur-rent evidence. Philosophical Transactions of the Royal Society B: Biological Sciences 372 (2017).

[16] T Garg, Ecosystems and human health: The local benefits of forest cover in indonesia. Journal of Environmental Economics and Management 98, 102271 (2019).

[17] S Bauhoff, J Busch, Does deforestation increase malaria prevalence? evidence from satel-lite data and health surveys. World Development 127, 104734 (2020).

[18] C Wing,K Simon, RA Bello-Gomez, Designing difference in difference studies: best practices for public health policy research. Annual Review of Public Health 39 (2018).

[19] BLS Jacobson, RJ Lalonde, DG Sullivan, Earnings Losses of Displaced Workers. American Economic Review 83, 685–709 (1993).

[20] MJ Bailey, A Goodman-Bacon, The War on Poverty’s Experiment in Public Medicine: Community Health Centers and the Mortality of Older Americans. American Economic Review 105, 1067–1104 (2015).

[21] A Goodman-Bacon, Public insurance and mortality: Evidence from medicaid implementation. Journal of Political Economy 126, 216–262 (2018).

[22] E Frank, The Effects of Bat Population Losses on Infant Mortality through Pesticide Use in the U.S. https://www.eyalfrank.com/research (2018) (year?).

[23] BA Jones, Forest-attacking Invasive Species and Infant Health: Evidence From the Invasive Emerald Ash Borer. Ecological Economics 154, 282–293 (2018).

[24] BA Jones, Infant health impacts of freshwater algal blooms: Evidence from an invasive species natural experimen. Journal of Environmental Economics and Management 96, 36–59 (2019).

[25] Pan American Health Organization, Health Information Platform for the Americas: Malaria Indicators (http://www.paho.org/data/index.php/en/ mnu-topics/indicadores-malaria-en.html) (2020) Accessed: 2020-1-16.

## References

[1] XP Song, et al., Global land change from 1982 to 2016. Nature 560, 639–643 (2018).

[2] C Kleinn,L Corrales, D Morales, Forest area in costa rica: a comparative study of tropical forest cover estimates over time. Environmental Monitoring and Assessment 73, 17–40 (2002).

[3] A Grainger, Rates of deforestation in the humid tropics: estimates and measurements. Geographical Journal 159 (1993).

[4] J Mummolo, E Peterson, Improving the Interpretation of Fixed Effects Regression Results. Political Science Research and Methods 6, 829–835 (2018).

[5] R Hijmans, High resolution monthly climate variables 1960-2018 (https://www.worldclim.org/data/monthlywth.html) (2019) Accessed: 2019-12-11.

[6] KR Lips, J Diffendorfer, JR Mendelson, MW Sears, Riding the wave: Reconciling the roles of disease and climate change in amphibian declines. PLoS Biology 6 (2008).

[7] R Puschendorf, F Bolanños, G Chaves, The amphibian chytrid fungus along an altitudinal transect before the first reported declines in costa rica. Biological Conservation 132, 136–142 (2006).

[8] FM Brem, KR Lips, Batrachochytrium dendrobatidis infection patterns among panamanian amphibian species, habitats and elevations during epizootic and enzootic stages. Diseases of Aquatic Organisms 81, 189–202 (2008).

[9] R Gagliardo, et al., The principles of rapid response for amphibian conservation, using the programmes in panama as an example. International Zoo Yearbook 42, 125–135 (2008).

[10] DC Woodhams, et al., Chytridiomycosis and amphibian population declines continue to spread eastward in panama. EcoHealth 5, 268–274 (2008).

[11] SJ Wright, MJ Samaniego, Historical, Demographic, and Economic Correlates of Land-Use Change in the Republic of Panama. Ecology and Society 13 (2008).

[12] CRU TS: Climate Research Unit Time Series v4.03 (http://dx.doi.org/10.5285/10d3e3640f004c578403419aac167d82) (2019).

[13] I Harris, PD Jones, TJ Osborn, DH Lister, Updated high-resolution grids of monthly climatic observations–the cru ts3.10 dataset. International Journal of Climatology 34, 623– 642 (2014).

[14] LE Hay, RL Wilby, GH Leavesley, A comparison of delta change and downscaled gcm scenarios for three mountainous basins in the united states 1. Journal of the American Water Resources Association 36, 387–397 (2000).

